# Impact of Patient Positioning on Hemodynamic Assessment: A Comparison of Supine and Upright Right Heart Catheterization in Pulmonary Hypertension and HFpEF

**DOI:** 10.1101/2025.02.24.25322825

**Authors:** Mohammed A. Chowdhury, Johanna Squires, David M. Systrom, Aaron B. Waxman

**Author notes:** **Corresponding Author** Aaron B Waxman, Director, Pulmonary Vascular Disease Program, Brigham and Women’s Hospital, Boston, MA.

## Abstract

**Background:** Right heart catheterization (RHC) is the gold standard for the objective diagnosis of heart failure with preserved ejection fraction (HFpEF) and pulmonary hypertension (PH). Typically performed in the supine position, RHC may not accurately reflect the hemodynamic changes that occur during upright activity. The effect of patient positioning on RHC measurements, especially in distinguishing between precapillary and postcapillary PH, has been underexplored. This study aims to compare hemodynamic measurements between supine and upright RHC and assess how patient positioning influences the diagnosis of PH.

**Methods:** We performed a retrospective observational study of patients who underwent both supine and upright RHC during invasive cardiopulmonary exercise testing (iCPET) at Brigham and Women’s Hospital between January 2015 and January 2024. Hemodynamic parameters were measured and compared between both positions.

**Results:** Compared to upright RHC, supine RHC showed consistently higher mean PAP (21 ± 8.5 mmHg vs. 16 ± 8 mmHg) and PCWP (12 ± 4.7 mmHg vs. 6.4 ± 4.9 mmHg). Supine RHC diagnosed significantly more cases of HFpEF (21.8% vs. 5.7%) and isolated postcapillary PH (10.9% vs. %), primarily due to overestimation of PCWP. Upright RHC had a higher specificity for detecting elevated PCWP (>15 mmHg) in patients with elevated NT-proBNP (92.8% vs. 74.3%), iCPET diagnosed exercise HFpEF (95.29% vs 80.92%), PCWP/CO>2 (98.27% vs 84.61%) and demonstrated stronger association with PCWP/CO>2 (OR 9.11, 95% CI 5-16) compared to supine RHC. The differences in mPAP and PCWP measurement between supine and upright RHC persisted regardless of sex, age, BMI, or severity of chronic lung disease.

**Conclusions:** Hemodynamic discrepancies between supine and upright RHC exist which can significantly affect the diagnosis of PH and HFpEF. Upright RHC provides more physiologically relevant data, potentially enhancing diagnostic accuracy and should be considered in the evaluation of patients with PH.

**Clinical Perspective:** *What is new?:*

- This study highlights significant differences in hemodynamic measurements of mean pulmonary artery pressure and pulmonary capillary wedge pressure between supine and upright RHC.
- The difference in supine and upright hemodynamics persists despite age, sex, BMI, FEV1 or smoking.
- Compared to upright RHC, supine RHC results in overestimation of PCWP resulting in misclassification of PH.

*What are the clinical implications?:*

- Utilizing upright RHC may enhance the accuracy of diagnosing PH and HFpEF, leading to better-targeted therapies and management strategies for patients.
- Improved diagnostic precision could reduce unnecessary treatments and hospitalizations associated with misdiagnosed heart failure and pulmonary hypertension.

## Introduction

Right heart catheterization (RHC) remains the gold standard for hemodynamic assessment and diagnosis of various forms of pulmonary hypertension (PH). Precise measurement of pulmonary artery and pulmonary capillary wedge pressures (PCWP) are critical for distinguishing precapillary from postcapillary PH. Conventionally, RHC is performed in the supine position due to convenient venous access, consistent zeroing of pressure transducers at a standardized anatomical reference point, and minimal variability of posture. In the supine position, there is an increase in ventricular preload due to enhanced venous return and reduced gravitational effects on blood pooling. These changes result in higher filling pressures^1^ and cardiac output^2^ potentially mischaracterizing the true hemodynamic status.^2-4^ This limitation is further compounded by patient-specific factors such as advanced age, obesity, chronic obstructive pulmonary disease, and sleep apnea, which independently affect venous return and intrathoracic pressures.^5-8^ Respirophasic variations during supine RHC can additionally complicate PCWP estimation. Campain et al. demonstrated that the extent of these variations is significantly influenced by age, right atrial pressure, body mass index, and percent predicted FEV1.^9^ While alternative maneuvers like fluid challenges and exercise RHC have been proposed to improve left-sided filling pressure evaluation, they predominantly share the inherent limitations of supine positioning.^10,11^ Hemodynamic assessment in the upright position offers a potential solution, promising a more physiologically accurate representation of cardiac hemodynamics.

Assessing hemodynamics in the upright position offers several advantages by accurately capturing the body’s physiological responses to gravity and posture. Evaluating hemodynamics in the upright position, whether standing or sitting, can offer valuable insights into blood volume and cardiac reserve, particularly in conditions such as heart failure or hypovolemia, where postural changes may exacerbate symptoms. In some cases, significant drops in blood pressure or increases in heart rate during the transition from supine to standing may go undetected if measurements are limited to the supine position. Patients with conditions such as postural tachycardia syndrome (POTS), vasovagal syncope, or preload failure may exhibit hemodynamic abnormalities that are apparent only in the upright position.^12-14^ In individuals with underlying lung disease or abnormal body habitus, interpreting hemodynamic data requires careful consideration of how these factors influence circulatory and respiratory dynamics when assessed in the supine position.^5,7-9,15,16^ Upright hemodynamic assessment enables a more comprehensive understanding of cardiovascular adaptation to gravity, posture, and autonomic regulation. Currently, comprehensive studies comparing hemodynamic assessments in supine versus upright positions are limited, and the extent of variation in these measurements, along with the clinical factors influencing these differences, remains unclear.

Adopting a more physiologically representative approach to hemodynamic assessment may enhance diagnostic accuracy, ultimately leading to improved patient management and outcomes. In this study, we evaluated the hemodynamic differences between supine and upright RHC and assessed the impact of patient positioning on the differentiation between precapillary and postcapillary pulmonary hypertension.

## Method

### Study design

We performed a retrospective, observational analysis comparing the hemodynamics of patients undergoing RHC in both supine and upright positions. The study population was drawn from the Brigham and Women’s Hospital Pulmonary Vascular Registry, a comprehensive database of patients who were evaluated for exertional intolerance of unknown etiology with invasive cardiopulmonary exercise testing (iCPET). All patients included in this analysis underwent sequential supine and upright RHC between January 1, 2015, and January 1, 2024. The study was approved by the Institutional Review Board at Brigham and Women’s Hospital. Informed consent was obtained from all patients prior to their enrollment in the registry.^17^

Hemodynamic assessment included pulmonary artery pressure, PCWP, cardiac output estimated by both direct and indirect Fick method, and systemic vascular resistance, in both supine and upright positions. Additional clinical data, such as demographics, PH classification, and relevant comorbidities, were also recorded.

### Right heart catheterization

All patients included in the registry underwent iCPET, which consisted of a resting supine RHC followed by an upright bicycle exercise stress test. A balloon-tipped, flow directed, triple-lumen, fluid-filled 7.5 Fr Swan-Ganz catheter (Baxter/Edwards, Deerfield, IL, USA) was inserted percutaneously under fluoroscopic and ultrasound guidance into the internal jugular vein into the internal jugular vein. Transducers were zeroed in the supine position based on the level of the right atrium, or phlebostatic axis^18^ and zeroed to atmospheric pressure. Following hemodynamic measurements, the pulmonary artery catheter was secured and a sterile dressing was applied. The patients were transported to the stress laboratory in order to undergo a symptom-limited incremental CPET using an upright cycle ergometer with a breath-by-breath assessment of gas exchange (ULTIMA CPX; Medical Graphics Corporation, St Paul, MN, USA).

Pulmonary and systemic hemodynamics were continuously and simultaneously monitored during rest and exercise (Xper Cardio Physiomonitoring System; Phillips, Melborne, FL, USA). Subjects were seated in the upright position on the cycle ergometer and the zero reference for the pressure transducers was 5 cm below the middle of the sternum. An electronic average of hemodynamic pressures over three respiratory cycles was used ^16^ The majority of RHC hemodynamic assessments were conducted by a single operator to reduce variability in measurement techniques and mitigate potential bias. Precapillary PH was defined as mPAP>20 mmHg, PCWP≤15 mmHg and PVR>2 WU. Combined Pre- and Post-capillary PH (Cpc-PH) was defined as mPAP>20 mmHg, PCWP>15 mmHg and PVR>2 WU.^19^ Isolated Post-capillary PH (Ipc-PH) was defined as mPAP>20 mmHg, PCWP>15 mmHg and PVR>2 WU. Heart failure with preserved ejection fraction (HFpEF) was defined as either Cpc-PH or Ipc-PH with a left ventricular ejection fraction ≥ 50%.

### Statistical analysis

The variables in the database were assessed for inconsistencies, outliers and missing data. Outliers were defined as having values more than 1.5 times the interquartile range and were removed to improve the data distribution profile. Inconsistent or unexpected values were also removed from the database. Missing data was imputed by mean or median based on the data distribution of the respective variables. Categorical variables were analyzed using Pearson’s Chi-Square test, while continuous variables were assessed using either the Kruskal-Wallis test or T-test, depending on the distribution of the data. Association between dependent and independent variables were assessed with binary logistic regression models.

## Result

A total of 1,307 patients were analyzed, with a mean age of 56 ± 16 years, including 66.2% females (n = 864) and 33.8% males (n = 443), with a mean body mass index (BMI) of 28.7 ± 7 kg/m^2^.

### Hemodynamic Measurements

#### Supine vs. Upright Right Heart Catheterization

Supine RHC recorded an average mean pulmonary artery pressure (mPAP) of 21 ± 8.5 mmHg, compared to 16 ± 8 mmHg in the upright position, with a mean difference of 5 ± 5.4 mmHg (p < 0.0001). In the supine position, 42% of patients were classified as having mPAP > 20 mmHg, versus 21% in the upright position. The average pulmonary capillary wedge pressure (PCWP) was 12 ± 4.7 mmHg supine and 6.4 ± 4.9 mmHg upright, showing a significant mean difference of 5.7 ± 4.6 mmHg (p < 0.0001). Supine RHC indicated PCWP > 15 mmHg in 21.8% of patients, compared to only 5.67% in the upright position. There was no significant difference in pulmonary vascular resistance between the supine and upright positions (see Fig 1).

**Figure 1:**
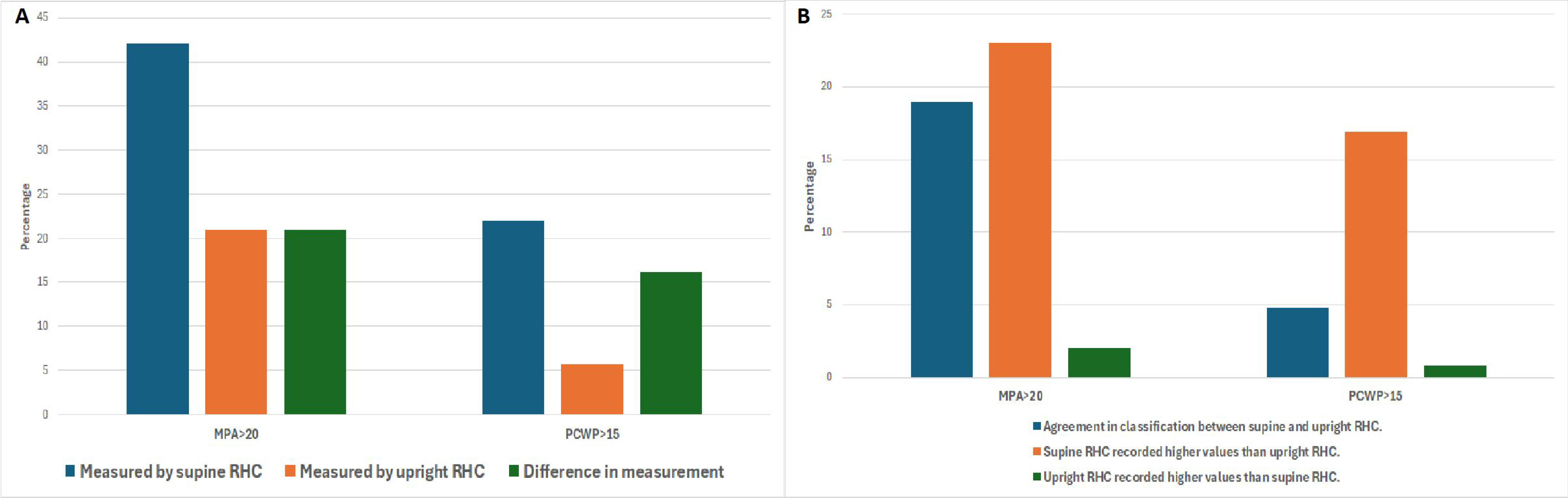
(A) Difference in hemodynamic measurement between supine and upright RHC (B) Differences in hemodynamic classification between supine and upright RHC.

#### Pulmonary Hypertension Classification

Precapillary PH was diagnosed in 13.4% via supine RHC and 12.3% via upright RHC. Cpc-PH was detected in 9.8% supine versus 3.52% upright. Ipc-PH was found in 10.87% supine compared to 1.84% upright. HFpEF was identified in 21.75% supine and 5.67% upright. Supine RHC generally overestimated mPAP and PCWP, leading to higher classification rates of HFpEF, Cpc-PH, and Ipc-PH (Table 1 and Fig 2).

**Table 1:** Differences in hemodynamic measurement and PH classification between supine and upright RHC.

|  | **Supine RHC** | **Upright RHC** | **Difference between**<br>**supine and upright**<br>**measurement** | **p value** |
| --- | --- | --- | --- | --- |
| Right atrial pressure (mmHg) | 6.8±3.5 | 2.7±3.5 | 4.1±3.8 | **<0.0001** |
| Mean pulmonary artery pressure (mmHg) | 21±8.5 | 16±8 | 5±5.4 | **<0.0001** |
| Pulmonary capillary wedge pressure (mmHg) | 12±4.7 | 6.4±4.9 | 5.7±4.6 | **<0.0001** |
| Pulmonary vascular resistance (WU) | 1.88±1.6 | 1.9±1.4 | -0.04±1.3 | 0.2 |
| Fick Cardiac output (L/min) | 5.3±1.2 | 5.6±1.8 | -0.3±2 | **<0.0001** |
| mPAP>20mmHg (n,%) | 42% (n=549) | 21% (n=275) | 21% | **<0.0001** |
| PCWP>15 mmHg (n,%) | 21.8% (n=284) | 5.67% (n=74) | 16.13% | **<0.0001** |
| Pulmonary arterial hypertension (n,%) | 13.4% (n=175) | 12.3% (n=161) | 1.1% | **<0.0001** |
| Combined pre and post capillary PH (n,%) | 9.8% (n=128) | 3.52% (n=46) | 6.28% | **<0.0001** |
| Isolated post capillary PH (n,%) | 10.87% (n=142) | 1.84% (n=24) | 9.03% | **<0.0001** |
| Heart failure with preserved ejection fraction (n,%) | 21.75% (n=284) | 5.67% (n=74) | 16.08% | **<0.0001** |

**Figure 2:**
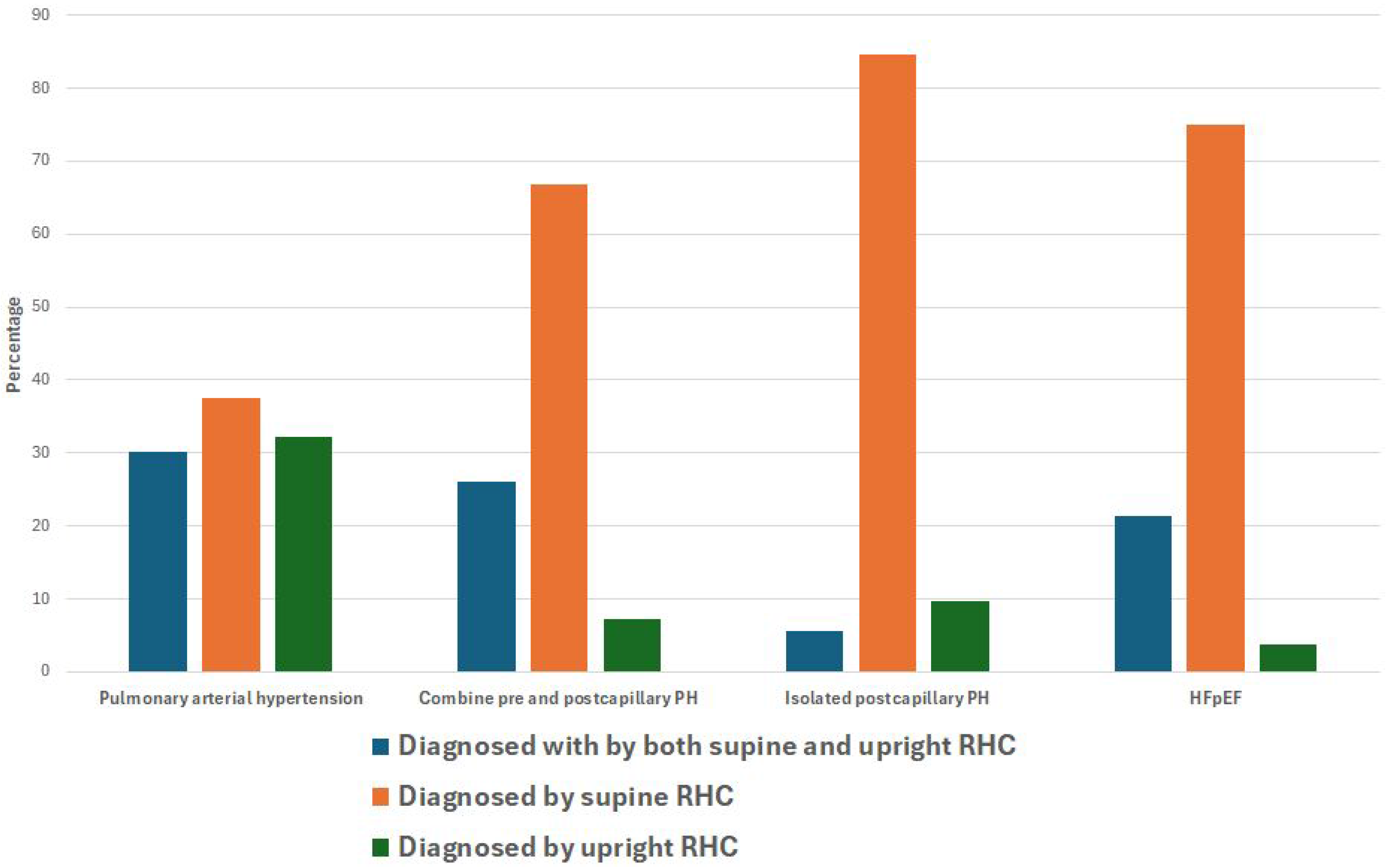
Differences in classification between supine and upright RHC.

#### Diagnostic Performance

Using upright RHC as the reference, a mPAP >20 mmHg, supine RHC had a sensitivity of 89.2% and specificity of 70.8%. For PCWP >15 mmHg, supine RHC had a sensitivity of 85.1% and specificity of 82.1%.

#### Influence of Physiological Factors

Males exhibited higher mPAP and PCWP than females in both positions, with supine measurements being consistently higher. Both mPAP and PCWP increased with advancing age, increasing BMI and worsening lung disease (Table 2), with supine values consistently higher than upright. Patients with history of smoking or CPAP/BiPAP use showed varied hemodynamic responses between positions, affecting mPAP less than PCWP (Table 3)

**Table 2:**
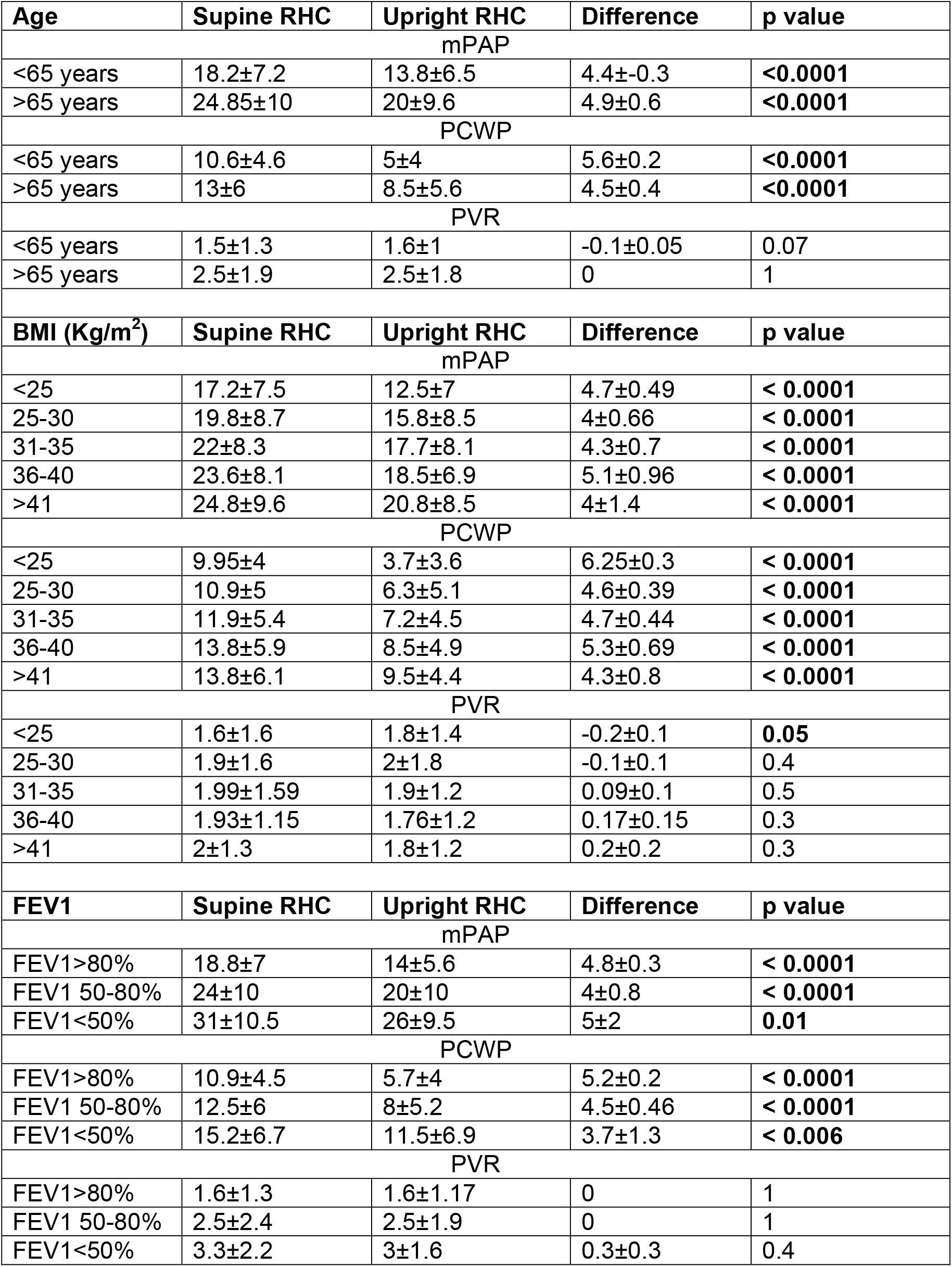
Differences in hemodynamic measurement between supine and upright RHC according to age, BMI and FEV1.

**Table 3:**
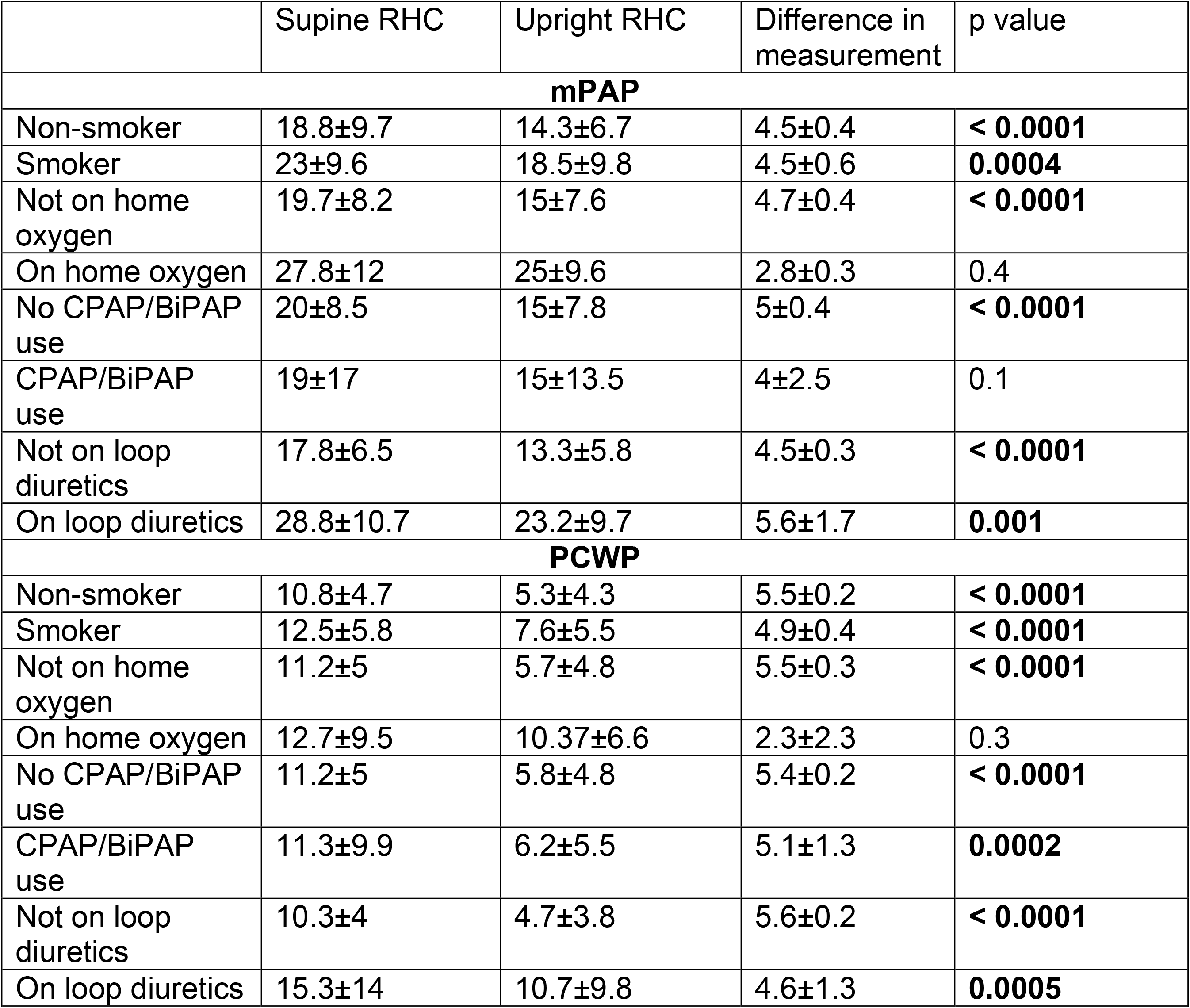
Influence of other factors on mPAP, PCWP and PVR stratified according to supine and upright RHC measurement.

|  | Supine RHC | Upright RHC | Difference in measurement | p value |
| --- | --- | --- | --- | --- |
| **mPAP** |  |  |  |  |
| Non-smoker | 18.8±9.7 | 14.3±6.7 | 4.5±0.4 | **< 0.0001** |
| Smoker | 23±9.6 | 18.5±9.8 | 4.5±0.6 | **0.0004** |
| Not on home oxygen | 19.7±8.2 | 15±7.6 | 4.7±0.4 | **< 0.0001** |
| On home oxygen | 27.8±12 | 25±9.6 | 2.8±0.3 | 0.4 |
| No CPAP/BiPAP use | 20±8.5 | 15±7.8 | 5±0.4 | **< 0.0001** |
| CPAP/BiPAP use | 19±17 | 15±13.5 | 4±2.5 | 0.1 |
| Not on loop diuretics | 17.8±6.5 | 13.3±5.8 | 4.5±0.3 | **< 0.0001** |
| On loop diuretics | 28.8±10.7 | 23.2±9.7 | 5.6±1.7 | **0.001** |
| **PCWP** |  |  |  |  |
| Non-smoker | 10.8±4.7 | 5.3±4.3 | 5.5±0.2 | **< 0.0001** |
| Smoker | 12.5±5.8 | 7.6±5.5 | 4.9±0.4 | **< 0.0001** |
| Not on home oxygen | 11.2±5 | 5.7±4.8 | 5.5±0.3 | **< 0.0001** |
| On home oxygen | 12.7±9.5 | 10.37±6.6 | 2.3±2.3 | 0.3 |
| No CPAP/BiPAP use | 11.2±5 | 5.8±4.8 | 5.4±0.2 | **< 0.0001** |
| CPAP/BiPAP use | 11.3±9.9 | 6.2±5.5 | 5.1±1.3 | **0.0002** |
| Not on loop diuretics | 10.3±4 | 4.7±3.8 | 5.6±0.2 | **< 0.0001** |
| On loop diuretics | 15.3±14 | 10.7±9.8 | 4.6±1.3 | **0.0005** |

#### Evaluation of PCWP measurement performance by supine and upright RHC

To evaluate the diagnostic performance of supine and upright RHC in detecting elevated left-sided filling pressures, we analyzed the association of PCWP >15 mmHg with abnormal NT-proBNP levels (>900 pg/mL), iCPET-diagnosed exercise HFpEF, defined by an incremental exercise PCWP/CO slope >2. Supine RHC demonstrated a sensitivity of 63.64% and specificity of 74.34% for detecting elevated PCWP in patients with abnormal NT-proBNP, while upright RHC exhibited lower sensitivity (27%) but higher specificity (92.8%). For detecting iCPET diagnosed exercise HFpEF, supine RHC showed a sensitivity of 55.79% and specificity of 80.92%, whereas upright RHC had a sensitivity of 17.89% and specificity of 95.29%.

When evaluating the association of PCWP >15 mmHg with PCWP/CO >2, supine RHC achieved a sensitivity of 55.29% and specificity of 84.61%, compared to a sensitivity of 26.44% and specificity of 98.27% for upright RHC. Logistic regression analysis revealed that elevated PCWP measured by upright RHC (OR 9.11, 95% CI 95% CI 5-16, p < 0.0001) had a stronger association with PCWP/CO >2 compared to supine RHC (OR 4.4, 95% CI 3-6, p < 0.0001). These findings indicate that while supine RHC tends to overestimate hemodynamic pressures, upright RHC offers greater specificity and may provide a more accurate assessment of left-sided filling pressures, particularly under physiological conditions relevant to pulmonary hypertension classification.

## Discussion

Our study reveals a significant discrepancy in the measurement of mPAP and PCWP between supine and upright RHC. Specifically, supine RHC tends to overestimate both mPAP and PCWP compared to upright RHC. This overestimation has critical implications for diagnosing various forms of PH, including precapillary PH, Cpc-PH, Ipc-PH, and HFpEF. It may misrepresent the hemodynamic changes that occur during upright exertion. Accurate diagnosis of PH relies on precise measurements of mPAP, PCWP, and pulmonary vascular resistance (PVR)^14^. Current diagnostic criteria, such as mPAP > 20 mmHg and PCWP < 15 mmHg, are predominantly based on supine RHC measurements. This raises concerns about potentially misclassifying patients with pulmonary arterial hypertension (PAH) as having Cpc-PH, which could prevent them from receiving targeted therapies essential for improving their quality of life and survival. Accurate classification is crucial because management strategies differ significantly. PAH therapies focus on the pulmonary vasculature, while Cpc-PH management addresses underlying left heart disease.^15^

The upright position reduces venous return, making it more sensitive for detecting preload failure. This can be particularly useful in evaluating patients with suspected HFpEF, including that which has been treated with diuretics and venodilators, where preload sensitivity is crucial. Upright measurements may better reflect the physiological conditions experienced during daily activities, as most patients complain of exertional intolerance in the upright position. This can help correlate symptoms like exertional dyspnea with hemodynamic changes more accurately.^20^ In the upright position, PVR tends to fall more significantly, especially at low levels of exercise. This can provide valuable information about pulmonary vascular function and its adaptation to different postures. Moreover, the upright position can reveal greater variability in the measurement of PCWP and other hemodynamic parameters, which might be masked in the supine position.^21^ This variability can be important for diagnosing conditions like pulmonary hypertension or assessing cardiac reserve. Overall, measuring hemodynamics in the upright position offers a complementary perspective to supine measurements, allowing for a more comprehensive assessment of cardiovascular function under different physiological stresses.

These effects are particularly pronounced in obese individuals, where elevated intra-abdominal pressure^15^ increases venous return, raising cardiac filling pressures. Additional challenges of supine RHC include respiratory variations that hinder accurate PCWP determination and pericardial restraint, which can result in right ventricular distension at the expense of left ventricular diastolic volume during increased preload.^9,22^ Moreover, supine measurements seldom reflect the hemodynamic changes that occur during physical activity in the upright position. Raina et al compared pulmonary pressures between baseline RHC and implantable hemodynamic monitors (IHM) and reported that, upon follow-up, approximately 48.8% of patients were identified as having pulmonary hypertension during ambulation based on IHM readings.^23^ Interestingly, these cases were not detected during the initial resting supine RHC highlighting the limitations and implications of our current practices. In contrast, upright RHC minimizes the above-mentioned confounding factors, providing hemodynamic measurements that more accurately reflect physiological conditions during daily activities. Our findings underscore the importance of incorporating upright hemodynamic assessments into clinical practice to improve diagnostic accuracy and ensure appropriate management of PH subtypes, ultimately optimizing patient outcomes.

The high specificity of upright RHC is particularly beneficial when evaluating patients suspected of having PH, as it reduces the likelihood of false positives in diagnosing elevated left-sided filling pressures. This minimizes the overestimation of PCWP and prevents the misclassification of patients with pulmonary arterial hypertension as having Cpc-PH. Consequently, this approach ensures that patients receive the correct diagnosis and appropriate care. Additionally, during exercise, both mPAP and PCWP increase significantly in both positions; however, the upright position more accurately reflects physiological conditions. Therefore, upright hemodynamic measurements are preferred during iCPET for a more accurate assessment.

Previous studies have similarly reported an overestimation of PCWP in the supine position compared to the upright position with a mean difference of approximately 5 mmHg.^2,4^ Our study has also shown a similar difference in PCWP measurement between the two positions. To the best of our knowledge, our study includes the largest sample of patients to date with simultaneous supine and upright hemodynamic measurements. Furthermore, we observed that mPAP and PCWP increase with advancing age, higher BMI, decreasing FEV1, and are also influenced by the patient’s sex, ongoing diuretic use, and smoking history. However, the differences between supine and upright measurements persist despite these factors, indicating that patient positioning significantly affects hemodynamic assessment even when adjusted for other confounding factors.

The limitations of this study include its retrospective design, which may introduce selection bias and unmeasured confounding variables. However, it is important to note that although the data analysis was conducted retrospectively, the data collection was prospective. Specifically, all patients were systematically scheduled to undergo a resting RHC followed by an upright hemodynamic assessment. The single-center nature of the research may limit the generalizability of the findings to other populations due to potential demographic homogeneity within the sample.

Our findings would suggest that it is time to consider an alternative approach to evaluation of patients with PH. Further studies are warranted to explore the feasibility, benefits, and outcomes of these proposed strategies.

## Data Availability

Dataset will be made available if required by the journal

